# Social Support, Traditional Care Practices, and Mental Health in Rural Ecuadorian Youth

**DOI:** 10.64898/2026.01.15.26344221

**Authors:** Rajendra P. Parajuli, Kristina Brandveen, Briana N.C. Chronister, Carlos F. Gould, Danilo Martinez, Dolores Lopez-Paredes, Jose R Suarez-Lopez

**Author notes:** These authors contributed equally to this work and share first authorship (Rajendra P. Parajuli, Kristina Brandveen). **Corresponding Author:** Jose Ricardo Suarez-Lopez, MD, PhD, Herbert Wertheim School of Public Health and Human Longevity Science, University of California San Diego (UCSD), 9500 Gillman Dr. San Diego, CA, USA. **RPP**. **KB**. **BNC**. **CG:**. **DM**. **DLP**. **JRS**.

## Abstract

Anxiety and depression account for a growing share of disability among adolescents and young adults globally, yet mental-health research and interventions focus primarily on clinical services, despite many in rural low- and middle-income settings relying on informal and culturally embedded systems of care. Using data from 488 emerging adults in a floriculture-intensive rural region of Ecuador, we quantified how ancestral medicine use and access to social support relate to anxiety and depression symptoms in a setting where formal mental-health care is limited. Anxiety and depressive symptoms were common (45% and 32%, respectively). Greater access to social support was strongly associated with lower depression scores and reduced odds of depressive symptoms, whereas associations with anxiety were weaker and more variable. In contrast, ancestral medicine use was associated with higher anxiety but showed no clear association with depression, indicating engagement with ancestral medicine as a care-seeking response to elevated psychological distress within a pluralistic care ecology rather than evidence of symptom alleviation. In contrast, access to social support was strongly associated with lower depression and reduced odds of depressive symptoms, with effects concentrated among males and among individuals reporting high relationship satisfaction and low loneliness. Pre-existing health conditions did not materially modify these associations. These patterns indicate that relationship quality governs whether access to social support translates into psychological protection, rather than social support operating as a uniformly protective exposure. These findings reframe youth mental health in rural agricultural settings as shaped less by engagement with specific therapeutic practices than by the structure and quality of social relationships, highlighting social support as a dominant correlate of resilience and ancestral medicine use as a marker of unmet mental-health need.

## 1. Main

Depression and anxiety affect over 300 million people worldwide and are leading contributors to disability, especially among adolescents and young adults ^1,2^. Despite the availability of effective psychological, pharmalolgical, and behavioral interventions, access to mental-health care remains limited in rural communities in low- and middle-income countries (LMICs), where shortages of trained providers, long travel distances, cost, stigma, and cultural misalignment between biomedical services and local expectations constrain care ^3–6^. As a result, mental health burdens among rural youth is substantial yet poorly captured by clinical systems.

In these settings, mental health is shaped not only by access to clinical services but by pluralistic care systems that include family and community social support as well as ancestral or traditional medicine (AM). Ancestral medicine encompasses diverse practices, including herbal treatments, spiritual cleansing, and ritual-based healing, that are widely used across Latin America, sub-Saharan Africa, and South and Southeast Asia, often serving as a first point of care where biomedical services are inaccessible or culturally incongruent ^4^ ^7–11^ ^12^ ^13^. Social support, from family, peers, and community members, has been consistently associated with better mental health ^14–17^. Social support may reduce exposure to stressors, enhance coping resources, and buffer physiological stress responses, thereby lowering the risk of common mental disorders.

However, much of the available evidence from LMICs comes from cross-sectional studies, often with limited adjustment for key confounders (e.g., socioeconomic status, pre-existing health conditions, loneliness, relationship satisfaction) and heterogeneous outcome measures, which constrain causal interpretation and cross-study comparability ^14,18–20^.

Ecuador provides a salient setting for studying how rural agricultural contexts shape mental health. Outside urban centers, depression and anxiety are frequently undiagnosed and untreated ^21^, even as national indicators show rising mental-health burden, particularly among women and agricultural workers. Recent national data show rising hospitalizations for depression-from 9.4 to 13.9 per 100 000 between 2015 and 2022, with women accounting for nearly two-thirds of cases ^22^. Burdens vary by geography, gender, and occupation ^24–27^: depression is estimated at 27% among women in Andean regions versus 35% on the coast ^23^, and moderate-to-severe depression reaches up to 63% among Andean highlander women ^28^. Ancestral medicine is widely practiced and legally recognized in Ecuador ^29,30^ under the country’s 1998 Constitution and 2006 *Ley Orgánica de Salud* ^31,32^. Despite its prevalence, and the coexistence of social networks that may offer psychosocial support, the mental-health benefits of AM and SS among adolescents and young adults in rural agricultural settings remain poorly understood.

Agricultural livelihoods introduce additional sources of psychological stress, including economic insecurity and chronic exposure to agrochemicals, which have been linked to neurobehavioral dysfunction and mood symptoms in rural Ecuador ^33–37^. Within this context, ancestral medicine and social support operate as central, socially legitimate components of care, yet their relationships to anxiety and depression among adolescents and young adults remain poorly characterized. Drawing on the Study of Secondary Exposures to Pesticides across Childhood, Adolescence, and Adulthood (ESPINA) longitudinal cohort in a floriculture-intensive rural region of Ecuador, we quantify the prevalence of anxiety and depression among emerging adults and examine how ancestral medicine use and access to social support relate to these outcomes. To clarify how pluralistic care systems map onto mental health burden in rural agricultural settings, we further assess whether these associations varied by sex, pre-existing health conditions, relationship satisfaction, and perceived loneliness - factors selected based on previous literature ^38–44^.

## 2. Materials and Methods

### 2.1 Study sample

ESPINA is a prospective cohort of children launched in 2008 in the agricultural canton of Pedro Moncayo, located in the Pichincha Province of Ecuador, to investigate the effects of pesticide exposure on human development.

Initially study participants were recruited in 2008 from the 2004 Survey of Access and Demand of Health Services (SADHS) in Pedro Moncayo County, which was conducted by the Fundación Cimas del Ecuador in collaboration with local community leaders and members.

Participants were also enrolled via community leaders, local councils, and word-of-mouth. To recruit a mixture of children with and without agricultural exposure, we enrolled 4-7 years-olds who met one of the following inclusion criteria: A) lived with a flower plantation worker for at least one year or B) never lived with an agricultural worker, stored pesticides at home, or had contact with pesticides. New participants were enrolled in 2016 and 2022, using the System of Local and Community Information (SILC) designed by Fundación Cimas del Ecuador, a geocoded database that includes information from the SADHS. Further information on data collection and recruitment has been described in detail in previous papers ^45,46^. In 2016 the cohort was expanded and two examinations were conducted: 330 participants in April (follow-up year [FUY] 8a) and 535 in July-October (FUY 8b), for a total of 554 participants examined in 2016 (311 in both time periods). Participants in 2016 were 12-17 years of age and included 238 participants examined in 2008 and 316 new volunteers. In July-September 2022 (FUY 14a), 505 participants were assessed, including 386 participants previously examined and 119 new participants. As in prior years, new participants were recruited using the SILC or by word of mouth. The present analysis includes 488 (96.6%) participants assessed in FUY 14a who had all relevant variables of interest.

### 2.2 Independent Variables

In FUY14a, ancestral medicine use was assessed by asking participants: “When you have a health problem, how often do you get treated by healers, yachags (local term traditional healer or shaman) or with ancestral medicine?” Responses were categorized as No use (1), Some use (2), and Always (3).

Access to social support was measured by asking: “When you have a problem, how often can you count on the help of others?” Response options included Never (1), Rarely (2), Sometimes (3), Frequently (4), and Always (5).

This and all other survey information was captured using a digital questionnaire (Qualtrics, LLC. Provo, UT) that each participant completed during the examination.

### 2.3 Outcome Variables

In FUY-14a, anxiety symptoms were measured using the Generalized Anxiety Disorder scale (GAD-7), a validated 7-item instrument that has been adapted for Spanish-speaking populations ^44^ and has previously been administered to Ecuadorian populations ^40^. Each item is rated on a 4-point Likert scale (0-3), yielding a total score range of 0-21. Symptom severity is categorized as minimal (0–4), mild (5–9), moderate (10–14), or severe (≥15) ^47^. Here, GAD-7 scores were used both as continuous variables and as binary indicators, with scores ≥5 classified as mild to severe anxiety.

Depressive symptoms were assessed using the Beck Depression Inventory-II (BDI-II), a 21-item self-report measure validated for use in Spanish-speaking populations ^38,39,41–43^. Items are scored on a 4-point Likert scale (0–3), producing a total score between 0 and 63. Symptom severity is classified as minimal (0–13), mild (14–19), moderate (20–28), or severe (≥29). BDI-II scores were treated as a continuous variable in linear models and a threshold of ≥14 was used to indicate mild to severe symptoms in logistic models ^48^.

### 2.4 Covariates

Sociodemographic and psychosocial covariates were collected in FUY14 using a digital questionnaire (Qualtrics, LLC. Provo, UT) completed by each participant during the examination. These included birthdate, gender, race/ethnicity, education, and monthly family income (in US$). Family monthly income (US$) was categorized as: 1 = <$100, 2 = $100–499, 3 = $500–999, and 4 = >$1000. For respondents that had missing family monthly income (n = 95), values were imputed using their reported 2016 monthly family income after adjusting for 8-year inflation (2016 income × 1.0862). If income was missing for 2016 and 2022 (n=19), the median 2016 income value ($580) was inflation-adjusted ($630) before categorization.

Preexisting health conditions were captured by asking participants to select among a list of commonly reported conditions for this age group, with an option to specify additional conditions not listed. Responses were coded as 0 (none) or 1 (some reported condition). Satisfaction with social relationships was measured by the question: “How satisfied are you with the way you relate to other people?” with responses including 1 (not at all satisfied), 2 (a bit satisfied), 3 (somewhat satisfied), 4 (mostly satisfied), and 5 (extremely satisfied). Perceived loneliness was measured by asking: “How often do you feel like you are alone?” with responses including 1 (rarely), 2 (occassionally), 3 (sometimes), 4 (often), and 5 (always).

In analyses, we binarized perceived satisfaction with social relationships and perceived loneliness by above and equal to or below the median: (i) somewhat, mostly, or extremely satisfied with social relationships vs. not at all or a bit satisfied and (ii) always feels alone vs. rarely, occasionally, sometimes, or often feels alone.

### 2.5 Statistical analysis

Baseline characteristics were summarized by AM use and SS level; group differences were tested using standard parametric and categorical tests. We fit separate regression models for AM and for SS. For continuous outcomes (GAD-7 and BDI-II), we used linear regression adjusting for age, gender, race, household monthly income, and years of formal education. For binary outcomes, we used logistic regression to estimate the odds of mild-to-severe anxiety (GAD-7 ≥ 5) and depression (BDI-II ≥ 14) with the same covariates. AM use was coded with no use as the reference category, and SS was dichotomized as frequently/always versus never/rarely/sometimes.

Effect modification by gender, preexisting health conditions, satisfaction with social relationships, and perceived loneliness was assessed by adding exposure-by-modifier interaction terms (AM×modifier or SS×modifier) in both the linear and logistic models. All models were then assessed in stratified analyses.

Statistical significance was defined as a p-value of less than 0.05. All the statistical analyses were carried out via R version 4.4.0 version 25.

### 2.6 Ethics Statement

The ESPINA study received ethical approval from the Institutional Review Boards of the University of Minnesota, the University of California San Diego, Universidad San Francisco de Quito, UTE University, and approved by the Ministry of Public Health of Ecuador. It also has the endorsement of the Commonwealth of Rural Parishes of Pedro Moncayo County. Informed consent was obtained from all adult participants. For minors, written assent was obtained along with parental or guardian permission for participation.

## 3. Results

### 3.1 Descriptive statistics

Participant characteristics are summarized in Table 1. The mean age was 20.29 years (SD=1.83), 51.2% were female, and 82% were Mestizo with the remaining 18% being Indigenous. More than one-third of participants reported symptoms of mild to severe anxiety (45% with GAD-7 ≥5) or depression (32% with BDI-II ≥14).

**Table 1:**
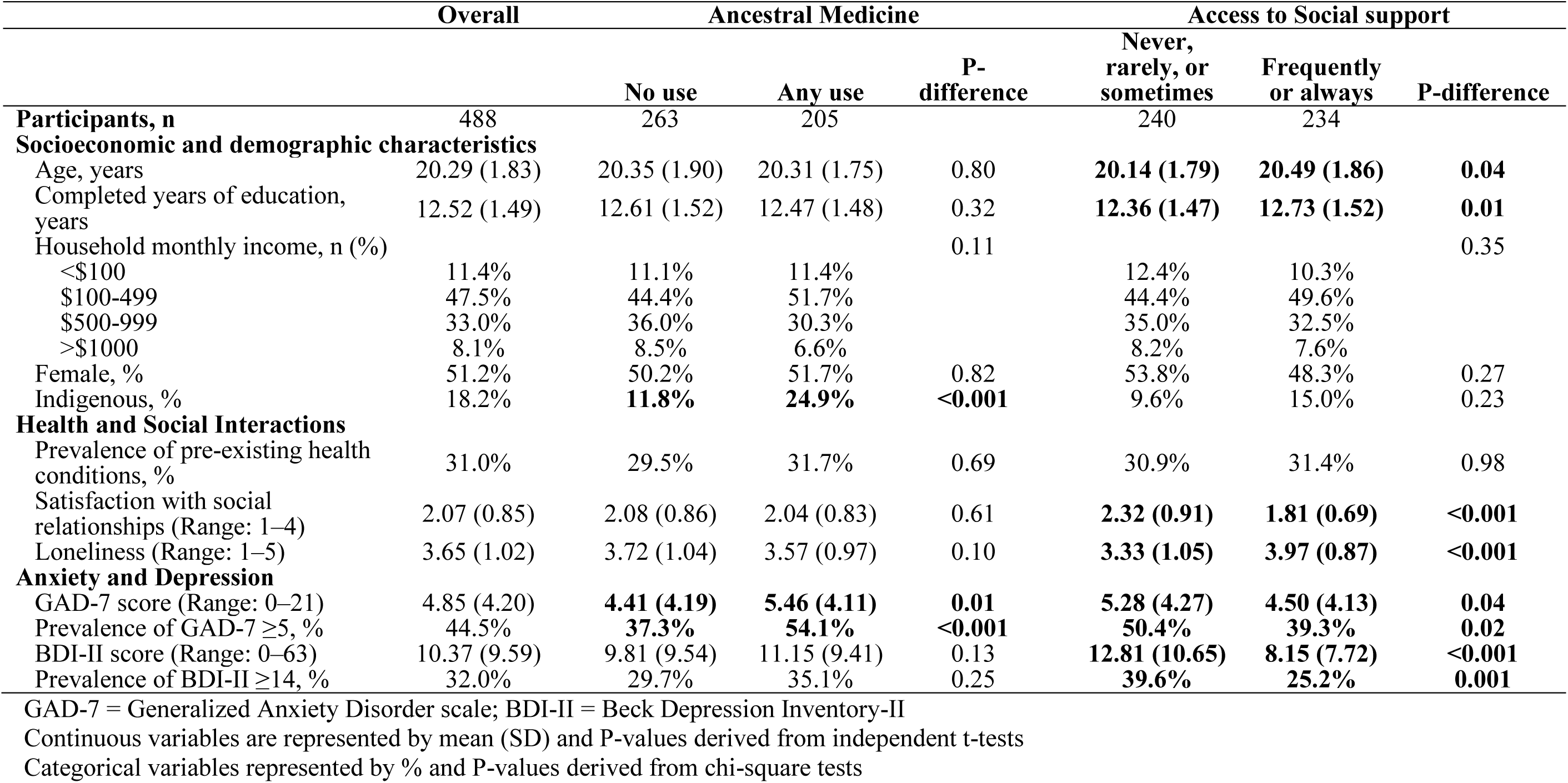
Descriptive characteristics of study participants in 2022, by use of ancestral medicine and social support.

Participants that used ancestral medicine had significantly higher GAD-7 scores and greater prevalence of mild to severe anxiety symptoms (p<0.01), though none of the sociodemographic or mental health characteristics differed between the two groups. Participants who frequently or always had access to social support were slightly older, had more years of formal education (p<0.01), lower BDI-II scores, and lower prevalence of mild to severe depression symptoms (p<0.01) than those with less access.

### 3.2 Use of ancestral medicine and mental health outcomes

After adjustment, ancestral medicine use was associated with higher anxiety scores (β = 1.00; 95% CI 0.23–1.76) and greater odds of mild to severe anxiety (OR = 2.00; 1.35–2.97; Table 2). Ancestral medicine use was also associated with higher depression scores (β = 1.34; –0.41 to 3.10), though this estimate was not statistically significant. Similarly, the odds of mild to severe depression were modestly higher among individuals reporting ancestral medicine use, but the association was not statistically significant (OR = 1.33; 0.87–2.01).

**Table 2.** Associations of ancestral medicine use and social support with anxiety outcomes among adolescents and young adults, including effect-modification and stratified analyses.

|  | **GAD-7 Score**<br>**Difference (95% CI)** | **Mild–Severe Anxiety**<br>**OR (95% CI)** |
| --- | --- | --- |
| **Ancestral medicine use (any vs none)** | **1.00 (0.23, 1.76) \*** | **2.00 (1.35, 2.97) \*** |
| **Effect modification by sex** |  |  |
| Interaction term | 0.68 (−0.83, 2.19) | 1.32 (0.60, 2.88) |
| Male | 0.66 (−0.37, 1.68) | 1.65 (0.92, 2.97) |
| Female | **1.30 (0.16, 2.45) \*** | **2.44 (1.39, 4.26) \*** |
| **Effect modification by pre-existing health conditions** |  |  |
| Interaction term | 0.04 (−1.59, 1.67) | 1.85 (0.75, 4.55) |
| No health conditions | **0.98 (0.07, 1.89) \*** | **1.78 (1.08, 2.91) \*** |
| Any health conditions | 1.23 (−0.25, 2.70) | **3.54 (1.56, 8.04) \*** |
| **Effect modification by perceived social relationship satisfaction** |  |  |
| Interaction term | −0.95 (−2.66, 0.75) | 0.94 (0.37, 2.38) |
| Low satisfaction | **1.22 (0.39, 2.04) \*** | **2.07 (1.30, 3.30) \*** |
| High satisfaction | 0.35 (−1.43, 2.14) | 2.17 (0.92, 5.16) |
| **Effect modification by perceived loneliness** |  |  |
| Interaction term | 0.31 (−1.46, 2.08) | 1.33 (0.46, 3.81), |
| Low loneliness | 0.65 (−0.27, 1.56) | **1.71 (1.09, 2.68) \*** |
| High loneliness | 0.72 (−0.49, 1.93) | 2.07 (0.66, 6.48) |
| **Social support (always/frequently vs less)** | **−0.70 (−1.46, 0.06)** | **0.67 (0.45, 0.98) \*** |
| **Effect modification by sex** |  |  |
| Interaction term | 1.41 (−0.12, 2.93) | **2.45 (1.11, 5.37) \*** |
| Male | **−1.42 (−2.44, −0.40) \*** | **0.39 (0.21, 0.71) \*** |
| Female | −0.06 (−1.21, 1.10) | 0.99 (0.58, 1.69) |
| **Effect modification by pre-existing health conditions** |  |  |
| Interaction term | 1.10 (−0.51, 2.71) | 2.20 (0.93, 5.19) |
| No health conditions | **−1.14 (−2.03, −0.24) \*** | **0.50 (0.30, 0.82) \*** |
| Any health conditions | −0.00 (−1.46, 1.46) | 1.18 (0.56, 2.48) |
| **Effect modification by perceived social relationship satisfaction** |  |  |
| Interaction term | −0.74 (−2.57, 1.09) | 0.78 (0.30, 2.03) |
| Low satisfaction | −0.11 (−0.94, 0.72) | 0.85 (0.54, 1.35) |
| High satisfaction | −0.89 (−2.81, 1.03) | 0.66 (0.28, 1.57) |
| **Effect modification by perceived loneliness** |  |  |
| Interaction term | 0.57 (−1.21, 2.35) | 2.37 (0.78, 7.25) |
| Low loneliness | −0.43 (−1.33, 0.47) | 0.69 (0.44, 1.07) |
| High loneliness | 0.52 (−0.70, 1.73) | 2.31 (0.67, 7.91) |

3 Mild to severe anxiety defined as GAD-7 $\geq 5$ ; \* $p < 0.05$
1 Sample size: N=488. Female (N=250) vs. Male (N=238); Any pre-existing health conditions (N=142) vs. No pre-existing health conditions (N=316); 2 High satisfaction with social relationships = somewhat, mostly, or extremely satisfied with social relationships (N=123) vs. low satisfaction = not at all 3 or a bit satisfied (N=352); High loneliness = always feels alone (N=116) vs. low loneliness = rarely, occasionally, sometimes, or often feels alone 4 (N=359). 5 Models adjusted for age in years, sex (excepting when stratified by sex), race, family income category, and formal education in years.

### 3.3 Access to social support and mental health outcomes

After adjustment, greater access to social support was associated with lower anxiety scores (β = –0.70; 95% CI –1.46 to 0.06) and lower odds of mild to severe anxiety (OR = 0.67; 0.45–0.98; Table 2). Social support was also associated with substantially lower depression scores (β = –4.52; –6.20 to –2.83, Table 3). Similarly, the odds of mild to severe depression were lower among individuals reporting greater social support (OR = 0.52; 0.35–0.79).

**Table 3.** Associations of ancestral medicine use and social support with depression outcomes among adolescents and young adults, including effect-modification and stratified analyses.

|  | **BDI-II Score**<br>**Difference (95% CI)** | **Mild–Severe Depression Score**<br>**OR (95% CI)** |
| --- | --- | --- |
| **Ancestral medicine use (any vs none)** | 1.34 (−0.41, 3.10) | 1.33 (0.87, 2.01) |
| **Effect modification by sex** |  |  |
| Interaction term | 1.63 (−1.83, 5.08) | 1.49 (0.64, 3.45) |
| Male | 0.46 (−1.90, 2.82) | 0.94 (0.48, 1.86) |
| Female | 2.16 (−0.43, 4.75) | 1.66 (0.95, 2.89) |
| **Effect modification by pre-existing health conditions** |  |  |
| Interaction term | 2.09 (−1.70, 5.88) | 1.75 (0.72, 4.26) |
| None | 0.89 (−1.24, 3.02) | 1.14 (0.67, 1.95) |
| Any | 3.33 (−0.03, 6.69) | **2.19 (1.01, 4.77) \*** |
| **Effect modification by perceived social relationship satisfaction** |  |  |
| Interaction term | −2.19 (−5.86, 1.48) | 0.58 (0.22, 1.48) |
| Low satisfaction | **2.03 (0.33, 3.73) \*** | 1.69 (0.99, 2.89) |
| High satisfaction | −1.33 (−5.24, 2.59) | 0.77 (0.31, 1.93) |
| **Effect modification by perceived loneliness** |  |  |
| Interaction term | 1.93 (−2.02, 5.89) | 1.66 (0.37, 7.38) |
| Low loneliness | 0.06 (−2.01, 2.13) | 1.07 (0.68, 1.70) |
| High loneliness | 1.82 (−0.60, 4.25) | 1.75 (0.35, 8.67) |
| **Social support**<br>**(always/frequently vs less)** | **−4.52 (−6.20, −2.83)\*\*** | **0.52 (0.35, 0.79) \*** |
| **Effect modification by sex** |  |  |
| Interaction term | 0.34 (−3.05, 3.72) | **1.05 (0.45, 2.47) \*** |
| Male | **−4.75 (−7.00, −2.50) \*** | **0.45 (0.23, 0.90) \*** |
| Female | **−4.58 (−7.14, −2.02) \*** | **0.48 (0.28, 0.84) \*** |
| **Effect modification by pre-existing health conditions** |  |  |
| Interaction term | 0.82 (−2.81, 4.46) | 1.89 (0.78, 4.54) |
| None | **−4.89 (−6.91, −2.86) \*** | **0.39 (0.22, 0.67) \*** |
| Any | **−4.58 (−7.87, −1.28) \*** | 0.69 (0.33, 1.43) |
| **Effect modification by perceived social relationship satisfaction** |  |  |
| Interaction term | −3.65 (−7.52, 0.22) | 0.79 (0.29, 2.14) |
| Low satisfaction | **−2.09 (−3.79, −0.40) \*** | 0.79 (0.46, 1.33) |
| High satisfaction | **−5.61 (−9.71, −1.50) \*** | 0.52 (0.20, 1.33) |
| **Effect modification by perceived loneliness** |  |  |
| Interaction term | **4.48 (0.61, 8.35) \*** | **1.90 (0.40, 8.98) \*** |
| Low loneliness | **−4.48 (−6.46, −2.49) \*** | **0.60 (0.38, 0.94) \*** |
| High loneliness | 0.54 (−1.90, 2.98) | 1.79 (0.31, 10.25) |

Mild to severe depression defined as BDI-II $\geq 14$ , \* $p < 0.05$ , \*\* $p < 0.001$
1 Sample Size: N=488. Female (N=250) vs. Male (N=238); Any pre-existing health conditions (N=142) vs. No pre-existing health conditions (N=316); 2 High satisfaction with social relationships = somewhat, mostly, or extremely satisfied with social relationships (N=123) vs. low satisfaction = not at all 3 or a bit satisfied (N=352); High loneliness = always feels alone (N=116) vs. low loneliness = rarely, occasionally, sometimes, or often feels alone 4 (N=359). 5 Models adjusted for age in years, sex (excepting when stratified by sex), ethnicity, family income category, and formal education in years.

### 3.4 Effect modification

#### 3.4.1 Gender

Associations between social support and anxiety differed by sex. Interaction terms for continuous anxiety and for the odds of mild to severe anxiety were positive (β_int = 1.41; –0.12 to 2.93; p=0.070; aOR_int = 2.45; 1.11–5.37; p=0.026; Table 3). In stratified analyses, social support was associated with markedly lower anxiety scores and lower odds of elevated anxiety among males, whereas estimates among females were close to the null. No evidence of sex modification was observed for depression outcomes or for associations with ancestral medicine use (Table 3).

#### 3.4.2 Pre-existing health conditions

Pre-existing health conditions modified associations between social support, ancestral medicine use, and mental health outcomes, although interaction terms were not statistically significant. In stratified analyses, higher social support was associated with lower anxiety scores and lower odds of mild-to-severe anxiety among participants without pre-existing health conditions (β = −1.14, 95% CI −2.03 to −0.24; aOR = 0.50, 0.30–0.82), but not among those with conditions (Table 2). Use of ancestral medicine was more strongly associated with anxiety among individuals with pre-existing health conditions (aOR = 3.54, 1.56–8.04) than among those without (aOR = 1.78, 1.08–2.91).

For depression, use of ancestral medicine was associated with higher odds of mild-to-severe depression among participants with pre-existing health conditions (aOR = 2.19, 1.01–4.77), whereas no statistically significant association was observed among those without conditions (Table 3). Higher social support was associated with lower depression scores regardless of health status, but lower odds of mild-to-severe depression were observed primarily among participants without pre-existing health conditions (aOR = 0.39, 0.22–0.67).

#### 3.4.3 Perceived satisfaction with social relationships

Perceived satisfaction with social relationships did not materially modify associations between social support and anxiety or depression, with interaction terms close to the null (Tables 2 and 3). Stratified analyses showed broadly protective associations of social support across satisfaction levels, with stronger reductions in depression scores among individuals reporting higher satisfaction. No evidence of effect modification was observed for ancestral medicine use by relationship satisfaction. However, among participants reporting low satisfaction with social relationships, ancestral medicine use was associated with higher anxiety scores and higher odds of mild-to-severe anxiety, as well as higher depression scores (Tables 2 and 3).

#### 3.4.4 Perceived loneliness

Loneliness did not significantly modify associations between social support and anxiety (Table 3). For depression outcomes, the interaction term for continuous scores was statistically significant, reflecting strong protective associations of social support among individuals with low loneliness and little to no association among those with high loneliness. We found no evidence that loneliness modified associations between ancestral medicine use and any mental health outcomes; interaction estimates were small and non-significant (Tables 2 and 3).

## 4. Discussion

In this study of emerging adults in a rural agricultural setting, we do not estimate the effects of specific mental-health treatments. Instead, we characterize how existing social and culturally embedded care systems map onto the distribution of anxiety and depression symptoms where formal mental-health services are limited. We find that access to social support is strongly associated with lower depressive symptoms, while ancestral medicine use tracks elevated anxiety, consistent with distress-driven care seeking rather than symptom reduction. Together, these findings suggest that youth mental health in rural settings is shaped less by engagement with particular therapeutic modalities than by the structure and quality of social relationships within pluralistic care systems.

The inverse association between SS and mental-health symptoms aligns with evidence that perceived support reduces depressive symptoms and buffers psychological distress across diverse settings ^16^. Although prior research in high-income countries has shown mixed associations with anxiety ^49^, our findings suggest benefits in this rural LMIC context. The modifying role of satisfaction with relationships and loneliness reinforce the importance of the quality of social relationships. Participants who reported high satisfaction with their relationships benefited the most from SS, while loneliness appeared to erode these benefits. These patterns mirror global evidence showing that loneliness is a strong, independent determinant of poor mental health, often superseding the effects of structural social networks ^50^. These findings also reflect dynamics common in rural and agricultural communities where geographic isolation, seasonal labor demands, and migration patterns can undermine social cohesion ^51^. The stronger protective effects observed among males in our sample may reflect gendered norms around emotional disclosure and help-seeking, as seen in other Latin American settings.

The positive association between AM use and anxiety is consistent with research showing that traditional and ancestral practices in Latin America are often used to address emotional or spiritual distress, particularly where formal mental-health services are limited, stigmatized, or culturally incongruent ^4,52,53^. AM use may also reflect broader structural vulnerabilities. Rural agricultural communities in Ecuador face high psychosocial burdens associated with poverty, food insecurity, climatic shocks, and occupational pesticide exposures—factors associated with anxiety and depression in prior research ^33,51^. In this context, AM practitioners often function as trusted, accessible points of care. These findings may also reflect broader historical transformations in Pedro Moncayo, where floriculture-driven economic shifts since the 1980s have altered gender roles, weakened intergenerational transmission of ancestral knowledge, and contributed to psychosocial stressors among youth and women ^54^. Although interaction tests were not statistically significant, AM users with high relationship satisfaction showed lower depression scores than those with low satisfaction, consistent with anthropological work on belonging and intergenerational continuity in Andean communities.

Emerging adulthood is a critical developmental period marked by heightened sensitivity to social and environmental stressors ^55^. In rural LMIC settings, this transition often occurs amid uncertainty about economic opportunities, environmental stress linked to agricultural production, and limited access to youth-focused mental-health care. The high prevalence of anxiety and depression observed here parallels reports of increasing psychosocial distress among youth in rural Latin America and underscores the need for contextually grounded approaches to mental-health support. Our findings contribute to this literature by showing that SS may function as a key determinant of resilience and that AM use reflects broader patterns of pluralistic care rather than a single, uniform practice.

Our study has several limitations. Its cross-sectional design precludes causal inference and limits our ability to disentangle whether AM use contributes to anxiety or whether anxiety motivates AM use. Mental-health measures and exposures were self-reported, raising the possibility of reporting bias influenced by stigma, social desirability, or differential cultural framing of symptoms. Data on the type, frequency, and purpose of AM use were limited; these are likely heterogeneous practices with diverse implications for mental health. Finally, data were collected during the COVID-19 recovery period, when residual psychosocial stressors may have influenced mental-health reporting ^56^.

Despite these limitations, the findings highlight the importance of strengthening social relationships as part of mental-health strategies for youth in rural Ecuador and suggest that AM plays a meaningful role in the broader landscape of care. Efforts to improve mental-health support should acknowledge and engage with AM practitioners, enhance SS networks, and expand culturally responsive services. Future research should employ longitudinal designs to clarify temporal pathways, use mixed methods to understand motivations and expectations surrounding AM use, and involve youth in the co-design of interventions that reflect their lived realities.

## 5. Conclusion

In a rural Ecuadorian cohort of emerging adults living in an intensive floriculture region, nearly half reported mild to severe anxiety and one in three reported mild to severe depression. Greater access to social support was strongly associated with lower anxiety and depression, whereas ancestral medicine use tracked higher anxiety—patterns consistent with protective effects of support and distress-driven care-seeking for AM. Associations varied by sex and by the perceived quality of social relationships. These findings support strengthening social-support components in community mental-health strategies and motivate longitudinal evaluation of how, when, and for whom AM contributes to mental-health care.

## Funding Source

The research presented in this manuscript was funded by the National Institute of Environmental Health Sciences (NIEHS) of the National Institutes of Health under Award Numbers R01ES025792, R01ES030378, and R21ES026084.

## Competing Interests

The authors have no relevant financial or non-financial interests to disclose.

## Author Contributions

Conceptualization: KB, JRS

Data Curation: BNC, DM, JRS

Methdology: KB, RPP, CFG, JRS

Formal analysis: KB, RPP, CFG

Investigation: BNC, DM, JRS

Writing – Original Draft: KB, RPP

Writing – Review & Editing: KB, RPP, BNC, DM, CFG, JRS

Supervision: CFG, JRS

Project Administration: JRS Funding Acquisition: JRS

## Data Availability

All data produced in the present study are available upon reasonable request to the authors

## Acknowledgments

We would like to express our sincere gratitude to all the participants who took part in this study. Our thanks also go to the ESPINA study team, Fundación Cimas del Ecuador, the Parish Governments of Pedro Moncayo County, the community members of Pedro Moncayo, and the Education District of Pichincha-Cayambe-Pedro Moncayo counties for their invaluable support and contributions to this project. The coauthors thank Dr. David Strong and Dr. Mateo Banegas for helpful comments.

## Notes

### Competing Interest Statement

The authors have declared no competing interest.

